# A Blinded Comparative Evaluation of Clinical and AI-Generated Responses to Otologic Patient Queries

**DOI:** 10.64898/2026.04.14.26350677

**Authors:** Sherifa Akinniyi, Ketan Jain-Poster, Emily Evangelista, Noriko Yoshikawa, Alexander Rivero

## Abstract

**Objective:** The objective of this study is to assess the quality, empathy, and readability of large language model (LLM) responses regarding otologic questions from patients as they compare to verified physician responses in other patient-driven forums. This study aims to predict the potential utility of LLMs in patient-centered communication.

**Study Design:** Comparative study

**Settings:** Internet

**Methods:** A sample of 49 otology-related questions posted on Reddit r/AskDocs^1^ between January 2020 and June 2025 were selected using search terms including “hearing loss,” “ear infection,” “tinnitus,” “ear pain,” and “vertigo.” Posts were retrieved using Reddit’s “Top” filter. Each question was answered by a verified doctor on Reddit and three AI LLMs (ChatGPT-4o, ClaudeAI, Google Gemini). Responses were scored by five evaluators.

**Results:** Common otologic concerns posed in patient questions were otalgia (38.7%), vertigo (28.6%), tinnitus (24.5%), hearing loss (22.4%), and aural fullness (20.4%). LLM responses were longer than physician responses (mean 145 vs 67 words; p < .05) and rated higher in quality (10.95 vs 9.58), empathy (7.26 vs 5.18), and readability (4.00 vs 3.73); (all p < .05). Evaluators correctly identified AI versus physician responses in 89.4% of cases with higher sensitivity for detecting physician responses (93.5%). By Flesch-Kincaid grade level, ChatGPT produced the most readable content (mean 7.25), while ClaudeAI responses were more complex (11.86; p < .05).

**Conclusion:** LLM responses received higher ratings in quality, empathy, and readability than those of physicians in response to a variety of otologic concerns. When appropriately implemented, such systems may enhance access to understandable otologic information and complement clinician-delivered care.

## INTRODUCTION

The recent surge in artificial intelligence (AI) has been applied to a wide variety of healthcare facets including diagnostic efficiency, drug development, clinical trial execution, risk stratification, and patient communication.^2^ Among the most visible AI applications are conversational chatbots powered by large language models (LLMs), such as ChatGPT, which generate natural language responses tailored to user queries, offering the potential to improve patient education, triaging, and healthcare engagement.^3^

In otologic care, AI has enabled advances in acoustic wearable technologies such as hearing aids and cochlear implants, as well as improvements in diagnostics through audiometry, imaging, and genetic profiling^4^. LLM-generated responses have shown potential to enhance clinical decision-making and referral practices^5^. In otology, LLMs have shown encouraging yet incomplete results. For example, ChatGPT has demonstrated success in identifying vestibular causes of dizziness in clinical vignettes, providing understandable and accurate information on cochlear implant recovery, offering reproducible and sound guidance on otosclerosis management, and aligning with expert opinions regarding monitoring after self-administered hearing tests.^6–9^

As patient reliance on digital health resources grows, it becomes essential to understand whether LLM responses can provide accurate, readable, and compassionate guidance^10^. A 2023 study evaluating LLM responses to publicly posted general medical questions found that blinded reviewers preferred LLM answers to physician responses nearly 78.6% of the time, rating them as higher in quality and empathy.^11^ Similar findings have been reported in oncology, where LLM outputs were comparable to or even exceeded physician responses in clarity and tone.^12^

Despite these advances, concerns remain regarding the accuracy, clinical appropriateness, and risk of misinformation in AI-generated medical content. Prior studies have typically examined a single condition or symptom, often relying on a single AI model, and none have directly compared LLM-generated responses with those of physicians. Accordingly, this study aimed to compare the quality, empathy, and readability of responses generated by multiple leading LLMs with physician responses from a public online forum addressing a broad range of otologic symptoms.

## METHODS

### Study Design and Data Collection

We conducted a comparative study using a sample of 49 otology-related questions posted on Reddit r/AskDocs between January 1, 2020, and June 30, 2025. Data was collected on June 20, 2025 using the search terms “hearing loss,” “ear infection,” “tinnitus,” “ear pain,” and “vertigo” in the “r/AskDocs” thread on Reddit. The frequency of each search term was collected and analyzed. Each question received responses from three LLMs (ChatGPT-4o, ClaudeAI Sonnet 4, Google Gemini) as well as from verified physician moderators on the Reddit r/AskDocs platform. Medical professionals are verified by submitting documentation (e.g., proof of licensure or institutional affiliation) to Reddit moderators, who review and confirm credentials before assigning a verified flair that identifies the user as a healthcare professional. There is no specialty-specific verification. LLMs were prompted with the following instruction: “You are a board-certified otolaryngologist providing responses at a sixth grade reading level to patients with the following ENT-related question. Use clear and medically accurate language. Please limit the length of the response to less than 100 words.” All responses were anonymized, blinded, and randomly ordered before evaluation. This review was deemed non-human subjects research and thus was exempt from approval by the institutional review board.

### Readability

Readability was assessed using the Flesch-Kincaid Grade Level (FKGL), which estimates the U.S. school grade level required for comprehension based on sentence length and word syllable count and was selected as the primary readability metric for interpretation. Supporting readability indices included the Automated Readability Index (ARI), which evaluates readability using characters per word and words per sentence, and the Gunning-Fog Index (GFI), which estimates years of formal education required to understand a text based on sentence length and the proportion of complex words. Lexical diversity and syntactic complexity were assessed using the Measures of Textual Lexical Diversity (MTLD) and Mean Dependency Distance (MDD), which reflects average syntactic distance between related words, respectively, as complementary linguistic measures.

### REDCap survey

Each evaluator (A.R., N.Y., K.J.-P., E.E., S.A.) independently rated each response for overall quality, empathy, and readability. Ratings were based on a 5-point Likert scale (1 = very poor, 2 = poor, 3 = acceptable, 4 = good, 5 = very good) and averaged to determine a total score (Table 4).

### Statistical Analysis

Descriptive statistics were calculated for word count, including means and 95% confidence intervals. Comparisons between LLM and physician responses were conducted using two-tailed Welch’s *t*-tests. One-way ANOVA and leave-one-out sensitivity analyses were performed to validate findings. Statistical analyses were performed using Python 3.12 in Google Colab. AI-assisted coding features available within the Google Colab environment were used to support coding and statistical analyses. All analyses were reviewed and verified by the authors. Statistical significance was defined as p < 0.05.

## RESULTS

Principal symptoms reflected in the question set include otalgia (38.7%), vertigo (28.6%), tinnitus (24.5%), hearing loss (22.4%), aural fullness (20.4%), otorrhea (6.1%), and other (10.2%) (Table 1 and 2). Each physician rater (n = 5) evaluated four responses for each of the 49 patient questions, resulting in 980 total response ratings. All LLM responses scored significantly higher than physician responses in mean measures of overall response quality (10.95 vs 9.58), empathy (7.26 vs 5.18), and readability (4.00 vs 3.73) (Table 1 and Figure 2). Post hoc leave-one-out analyses further demonstrated that pooled LLM responses continued to significantly outperform physician responses across all dimensions, even when any single model (ChatGPT-4o, ClaudeAI Sonnet 4, or Google Gemini) was excluded (all p < .01) (Table 3). Evaluators accurately identified the response author type in 89.4% of cases, demonstrating greater sensitivity for recognizing physician responses in 93.5% of cases. Mean word counts of all LLM responses were larger than physician responses (mean, 145 [95% CI, 108.38-183.06]) (Table 1 and Figure 3). With respect to FKGL, ChatGPT produced the most readable responses (mean, 7.25 [95% CI, 6.83-7.68]), corresponding to approximately a seventh grade reading level, whereas ClaudeAI Sonnet 4 responses were significantly more complex and were written at a high school to college level (mean, 11.86 [95% CI, 10.91-12.80]). Physician (mean, 8.12 [95% CI, 7.12-9.13]) and Google Gemini (mean, 8.62 [95% CI, 8.07-9.16]) responses were intermediate, averaging eighth- to ninth-grade readability (Figure 1). Supporting readability metrics such as ARI and GFI, along with lexical and syntactic complexity measures including MTLD and MDD, demonstrated similar relative patterns across responses and are included in Table 1 and Figure 4.

**FIGURE 1.**
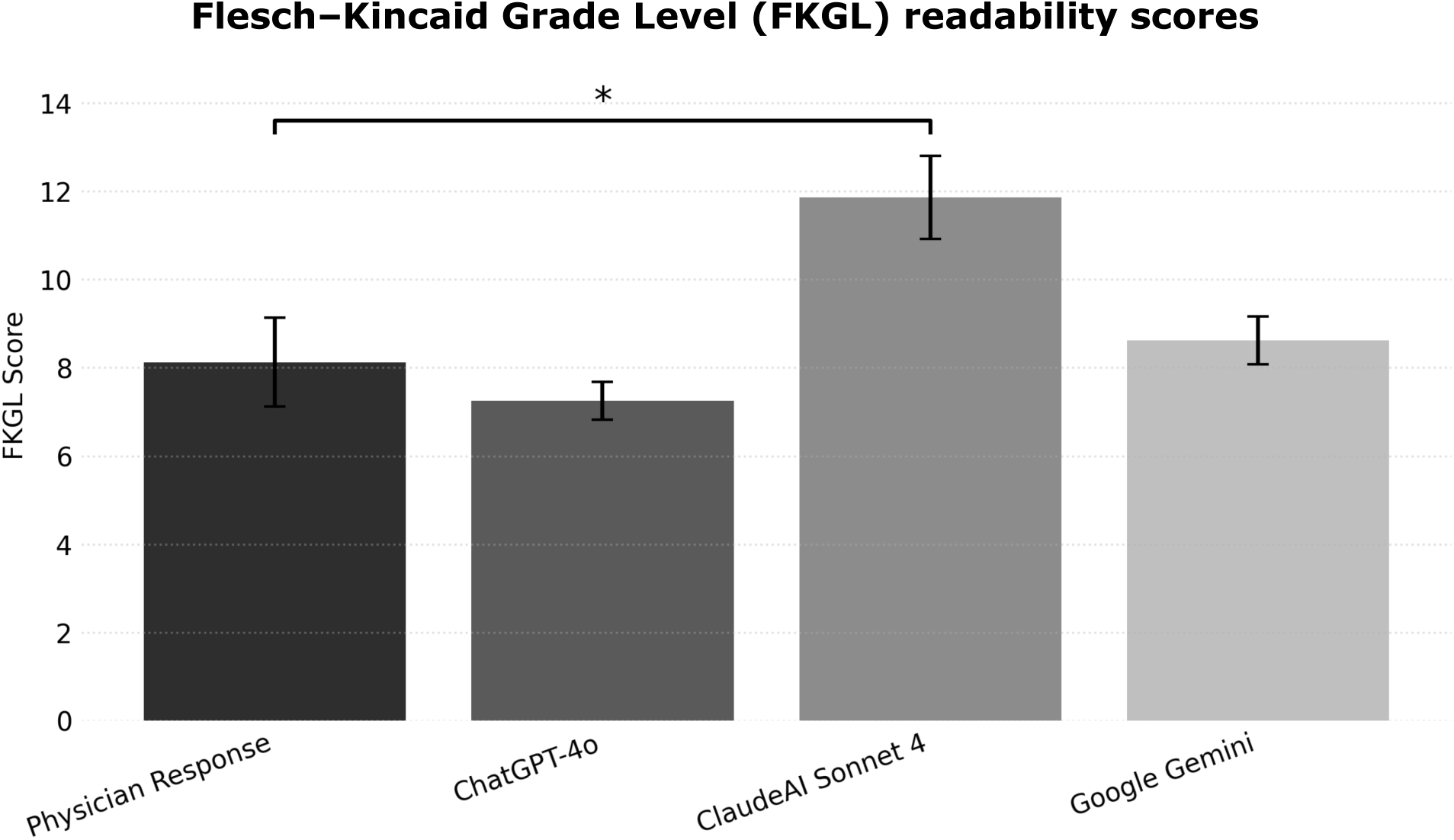
Mean Flesch–Kincaid Grade Level (FKGL) readability scores with 95% confidence intervals for physician responses and large language model responses. * Mean differences are statistically significant between LLM and physician response (p < 0.05).

**FIGURE 2.**
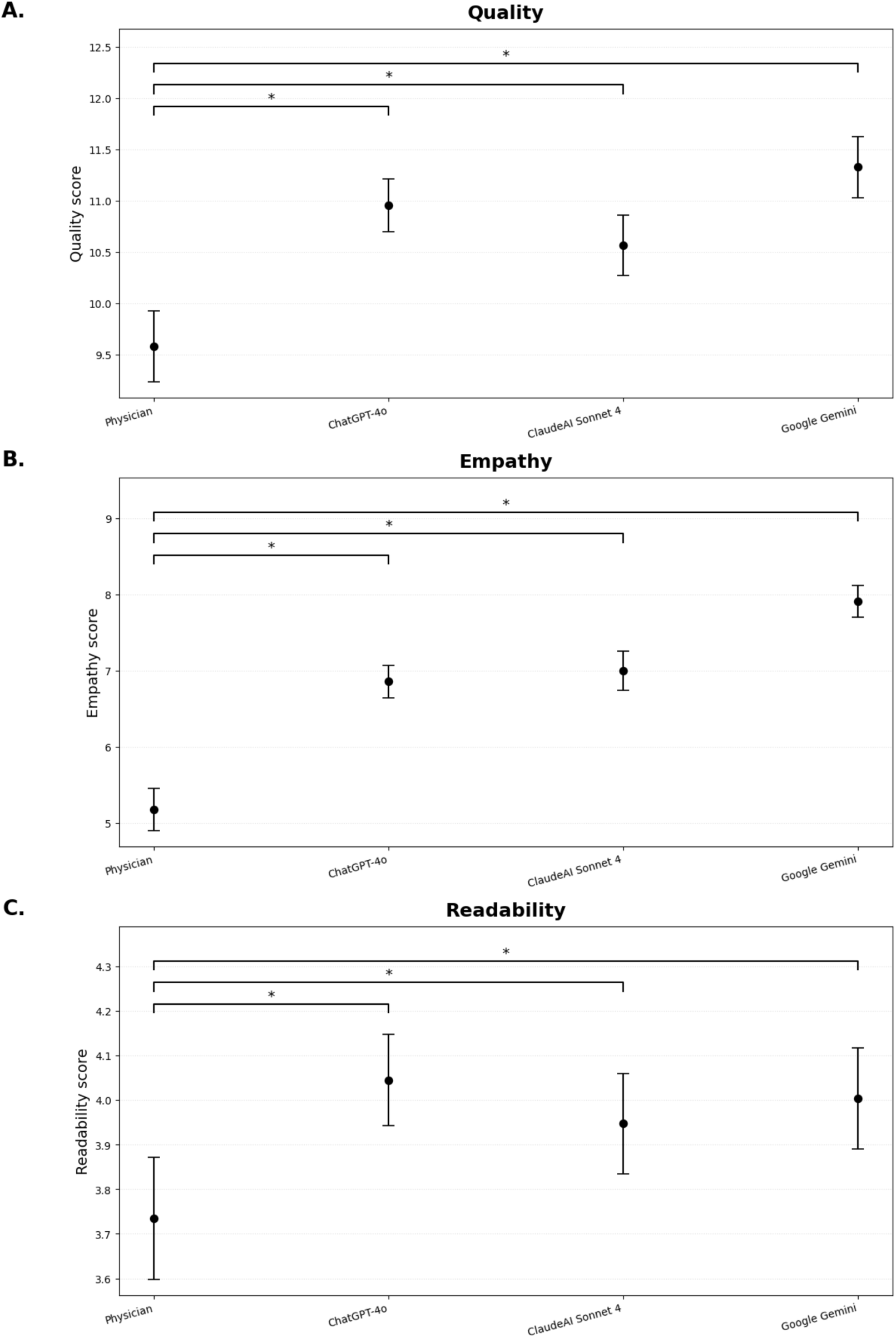
Distribution of overall quality, empathy, and readability scores for physician and large language model responses to patient questions. * Mean differences are statistically significant between LLM and physician response (p < 0.05).

**FIGURE 3.**
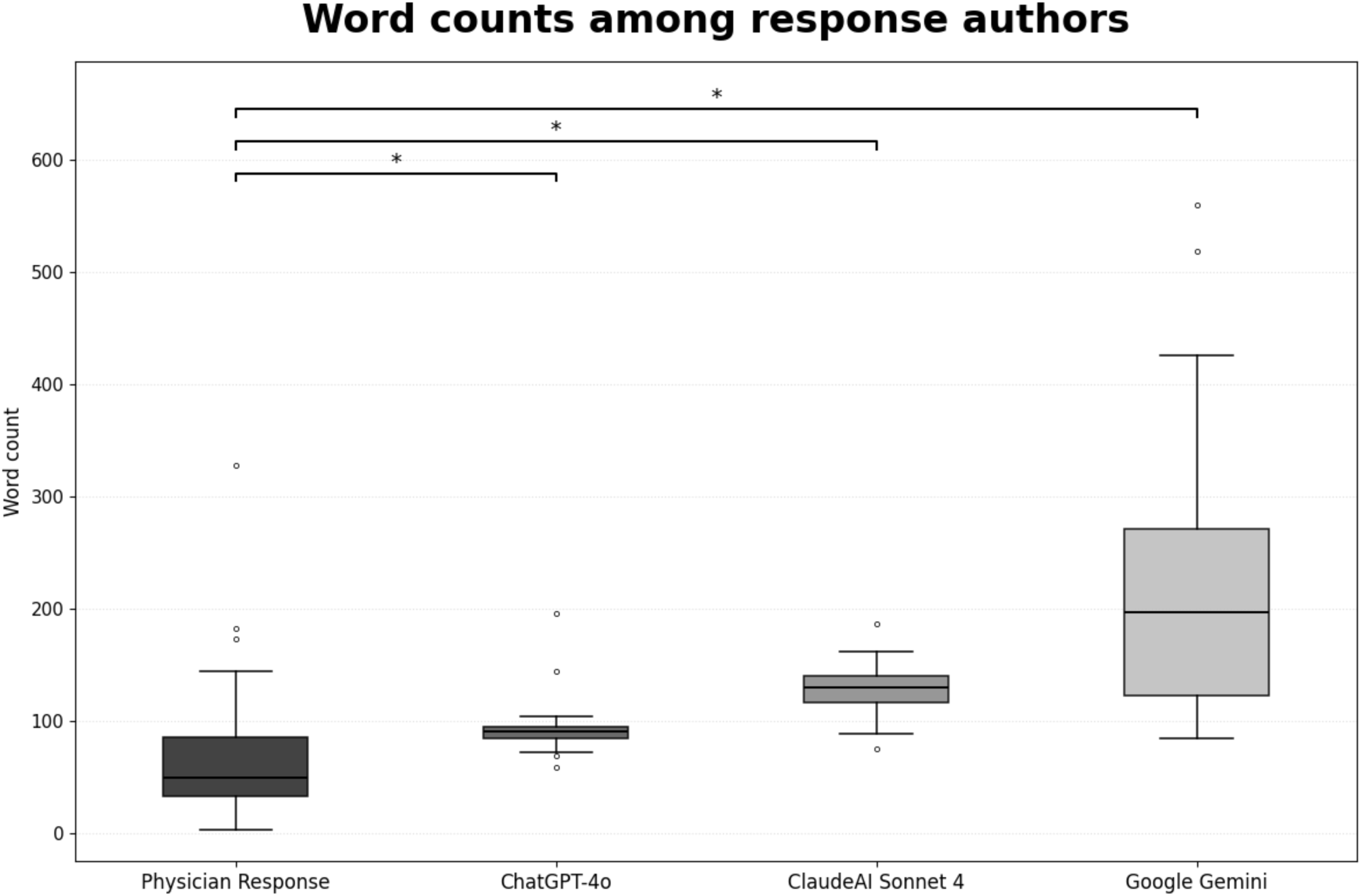
Mean word counts with 95% confidence intervals for physician responses and large language model responses. * Mean differences are statistically significant between LLM and physician response (p < 0.05).

**FIGURE 4.**
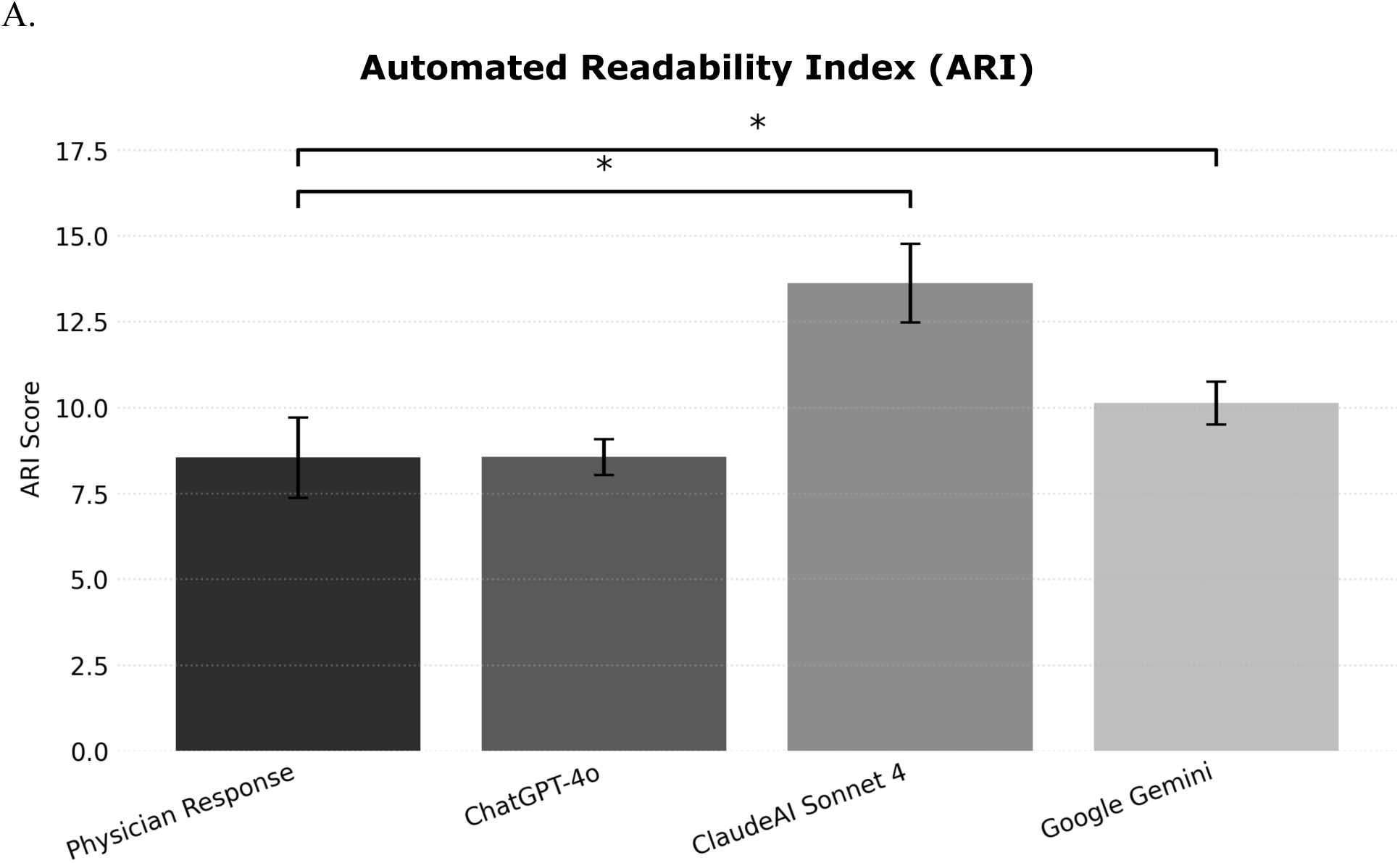
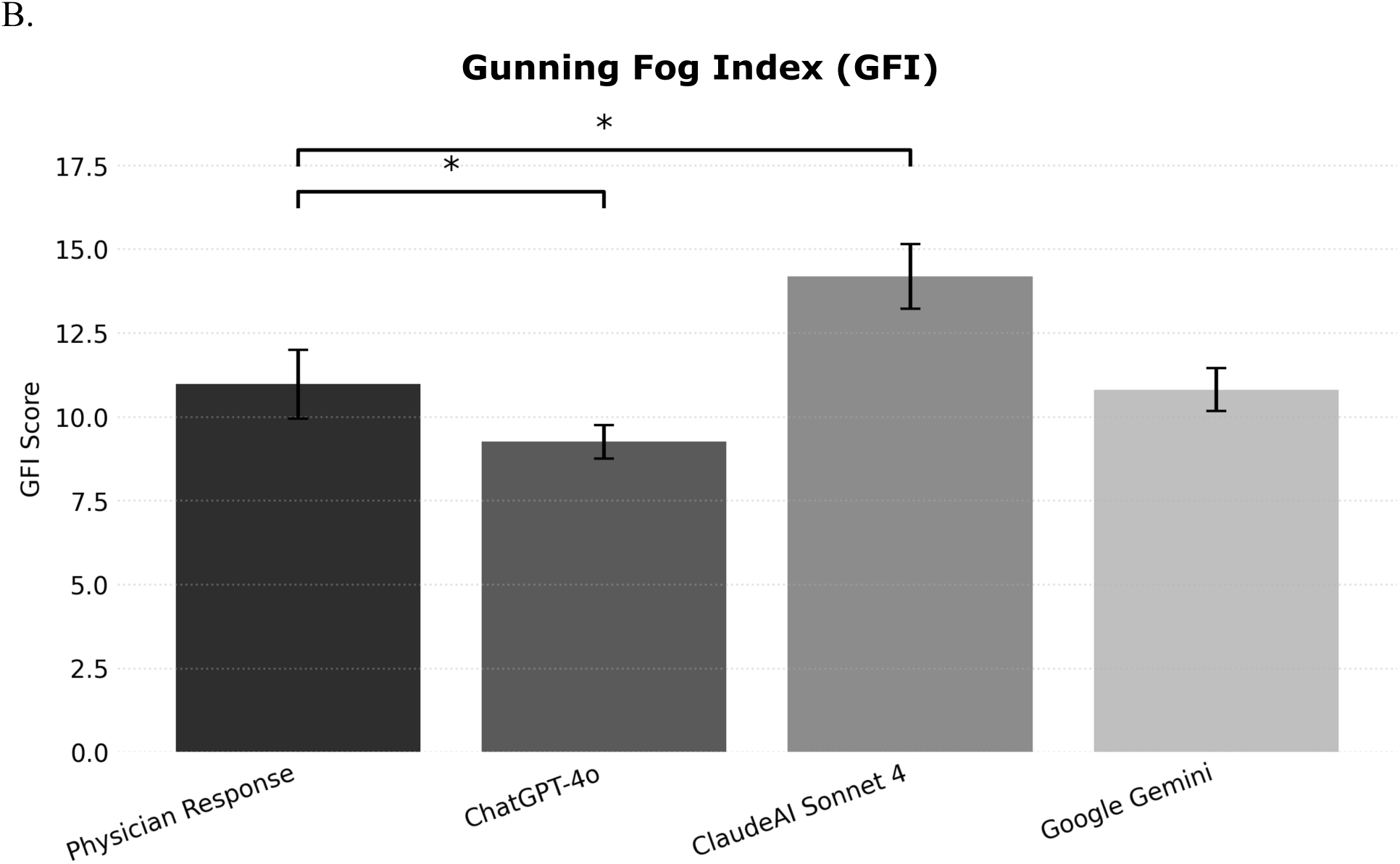
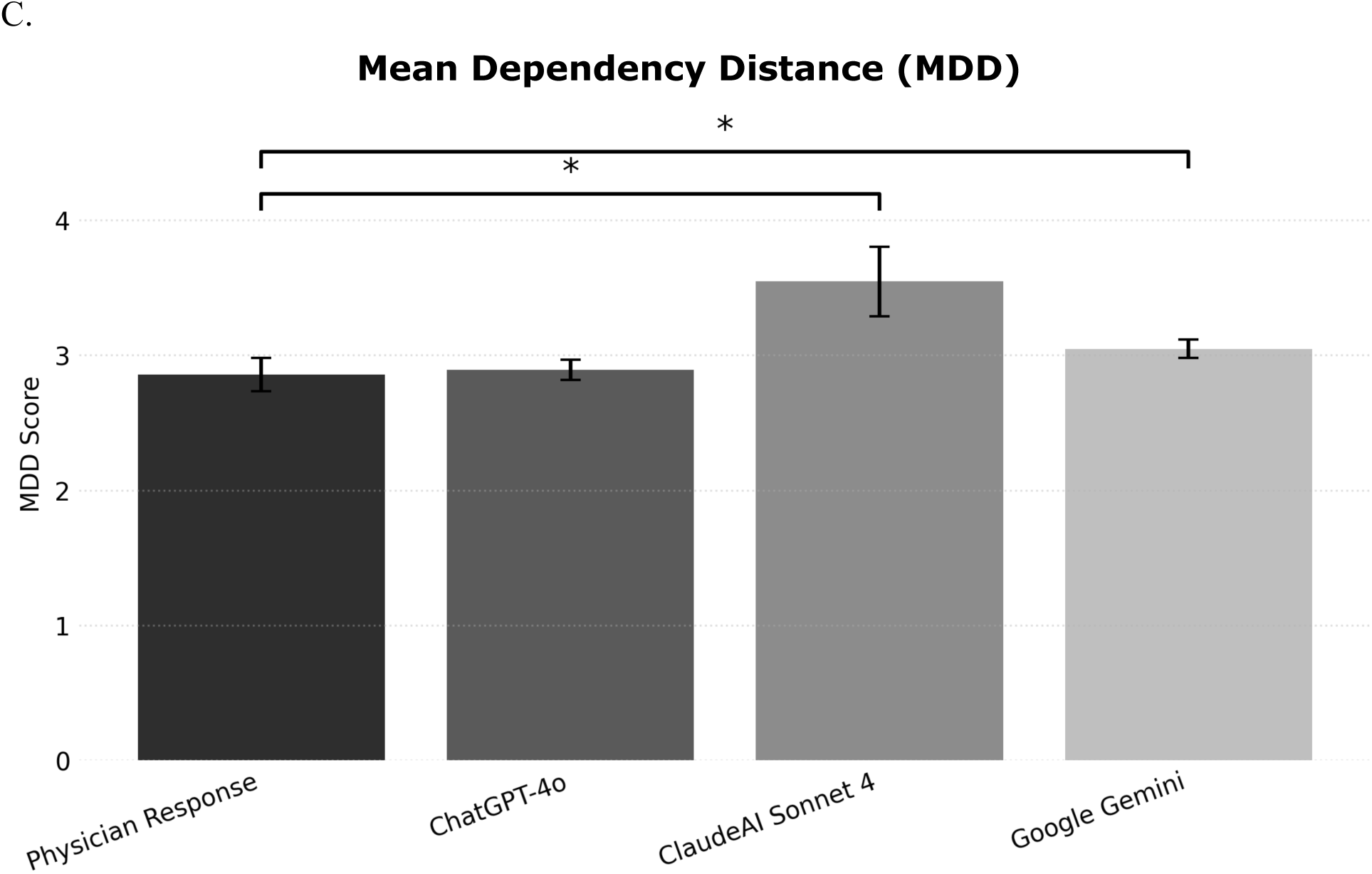
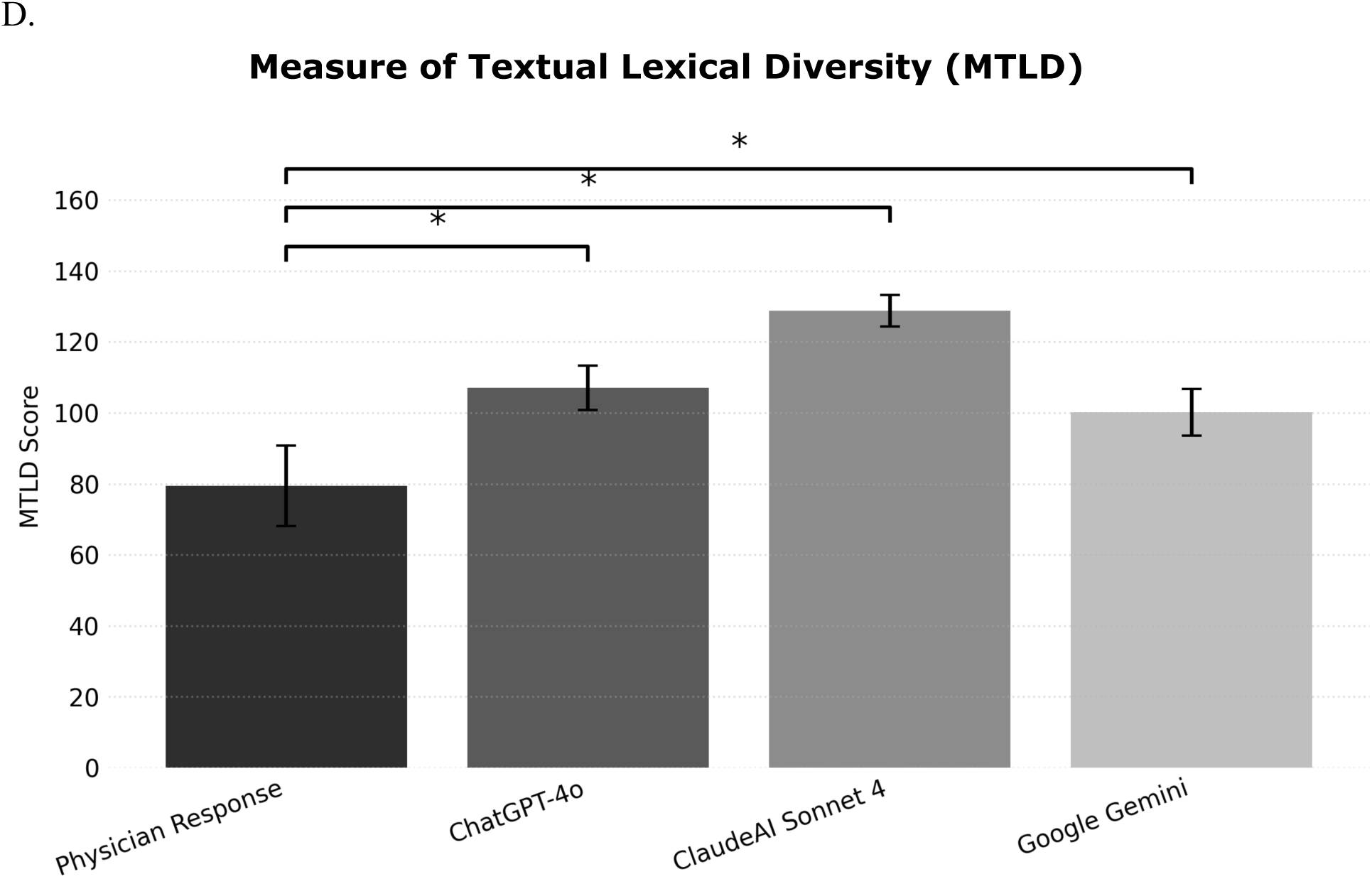
Readability, syntactic complexity, and lexical diversity metrics for physician responses and large language model (LLM)–generated responses to patient questions. (A) Automated Readability Index (ARI); (B) Gunning Fog Index (GFI); (C) Mean Dependency Distance (MDD); (D) Measure of Textual Lexical Diversity (MTLD). * Mean differences are statistically significant between LLM and physician response (p < 0.05).

**TABLE 1.** Mean Scores of quality, empathy, readability, syntactic complexity, and lexical diversity of physician responses and LLM responses to patient questions.

|  |  |  |  |  |
| --- | --- | --- | --- | --- |
|  | **Response, mean (95% CI)** |  |  |  |
| **Evaluation category and metric** | Physician | ChatGPT-4o<br>(Mean, 95% CI) | ClaudeAI Sonnet 4<br>(Mean, 95% CI) | Google Gemini<br>(Mean, 95% CI) |
| **Rater-evaluated quality score** |  |  |  |  |
| **Medical accuracy** | 3.27 (3.14-3.39) | 3.57 (3.46-3.67)<sup>a</sup> | 3.32 (3.19-3.45) | 3.76 (3.63-3.88)<sup>a</sup> |
| **Completeness** | 2.91 (2.77-3.05) | 3.60 (3.50-3.70)<sup>a</sup> | 3.73 (3.62-3.85)<sup>a</sup> | 4.00 (3.89-4.11)<sup>a</sup> |
| **Focus** | 3.40 (3.27-3.53) | 3.79 (3.70-3.88)<sup>a</sup> | 3.52 (3.41-3.62) | 3.60 (3.48-3.72)<sup>a</sup> |
| **Quality overall score** | 9.58 (9.24-9.92) | 10.96 (10.70-11.21)<sup>a</sup> | 10.56 (10.27-10.86)<sup>a</sup> | 11.33 (11.03-11.62)<sup>a</sup> |
| **Rater-evaluated empathy score** |  |  |  |  |
| **Emotional empathy** | 2.47 (2.34-2.61) | 3.38 (3.26-3.49)<sup>a</sup> | 3.46 (3.33-3.59)<sup>a</sup> | 4.01 (3.90-4.11)<sup>a</sup> |
| **Cognitive empathy** | 2.68 (2.54-2.83) | 3.49 (3.38-3.61)<sup>a</sup> | 3.53 (3.40-3.66)<sup>a</sup> | 3.91 (3.79-4.02)<sup>a</sup> |
| **Empathy overall score** | 5.18 (4.91-5.45) | 6.86 (6.64-7.07)<sup>a</sup> | 7.00 (6.75-7.25)<sup>a</sup> | 7.91 (7.71-8.12)<sup>a</sup> |
| **Rater-evaluated readability score** |  |  |  |  |
| **Readability overall score** | 3.73 (3.60-3.87) | 4.04 (3.94-4.15)<sup>a</sup> | 3.95 (3.84-4.06)<sup>a</sup> | 4.00 (3.89-4.12)<sup>a</sup> |
| **Word count** | 67 (51-83) | 92 (86-97)<sup>a</sup> | 129 (123-135)<sup>a</sup> | 216 (183-249)<sup>a</sup> |
| **Readability metrics** |  |  |  |  |
| **Flesch-Kincaid Grade Level (FKGL)** | 8.12 (7.12-9.13) | 7.25 (6.83-7.68) | 11.86 (10.91-12.80)<sup>a</sup> | 8.62 (8.07-9.16) |
| **Automated Readability Index (ARI)** | 8.54 (7.37-9.71) | 8.56 (8.04-9.08) | 13.62 (12.47-14.77)<sup>a</sup> | 10.13 (9.51-10.76)<sup>a</sup> |
| **Gunning Fog Index (GFI)** | 10.97 (9.94-12.00) | 9.25 (8.75-9.76)<sup>a</sup> | 14.19 (13.22-15.15)<sup>a</sup> | 10.81 (10.16-11.45) |
| **Syntactic Complexity** |  |  |  |  |
| **Mean Dependency Distance (MDD)** | 2.86 (2.73-2.98) | 2.89 (2.82-2.97) | 3.55 (3.29-3.80)<sup>a</sup> | 3.05 (2.98-3.11)<sup>a</sup> |
| **Lexical Diversity** |  |  |  |  |
| **Measure of Textual Lexical Diversity (MTLD)** | 79.47 (68.11-90.83) | 107.03 (100.75-113.31)<sup>a</sup> | 128.83 (124.40-133.25)<sup>a</sup> | 100.09 (93.53-106.66)<sup>a</sup> |

<sup>a</sup> Mean differences are statistically significant between LLM and physician response (p<0.05).
CI, confidence interval.

**TABLE 2.** Breakdown by Otologic Symptom.

| **TABLE 2. Breakdown by Otologic Symptom** |  |
| --- | --- |
| **Symptom** | **Count (% of questions)** |
| Otalgia | 19 (38.7%) |
| Vertigo | 14 (28.6%) |
| Tinnitus | 12 (24.5%) |
| Hearing loss | 11 (22.4%) |
| Aural fullness | 10 (20.4 %) |
| Otorrhea | 3 (6.1%) |
| Other<sup>a</sup> | 5 (10.2%) |

<sup>a</sup> Patient questions indicating self-diagnosed ear infections were categorized as “Other” for analysis.

**TABLE 3.** ANOVA and Leave-one-out analysis.

| **TABLE 3. ANOVA and Leave-one-out analysis** |  |  |  |  |  |
| --- | --- | --- | --- | --- | --- |
| **Dimension** | **ANOVA F(df)** | **p-value** | **Leave-one-out**<br>**(ChatGPT-4o)** | **Leave-one-out**<br>**(ClaudeAI Sonnet 4)** | **Leave-one-out**<br>**(Google Gemini)** |
| Overall<br>Quality | F(3,976)=24.20 | <0.001 | t=6.65, p<0.001 | t=7.75, p<0.001 | t=5.86, p<0.001 |
| Overall<br>Empathy | F(3,976)=87.67 | <0.001 | t=13.92, p<0.001 | t=13.76, p<0.001 | t=10.73, p<0.001 |
| Overall<br>Readability | F(3,976)=5.42 | 0.001 | t=3.01, p=0.003 | t=3.66, p<0.001 | t=3.30, p=0.001 |

**TABLE 4.** Definitions of component and overall measures of quality, empathy, and readability LLM response evaluation by physician.

| **TABLE 4.** Definitions of component and overall measures of quality, empathy, and readability LLM response evaluation by physicians. |  |  |  |
| --- | --- | --- | --- |
| **Metric** | **Quality** | **Empathy** | **Readability** |
| Medical Accuracy: How well the response is supported by evidence-based, peer-reviewed research and reflects a conclusion that a general physician would make | ✓ |  |  |
| Completeness: How well each response answers all parts of the patient question | ✓ |  |  |
| Focus: How well each response includes only relevant information to the patient question | ✓ |  |  |
| Overall Quality Score: Overall quality score based on the rater's ranked preference of responses | ✓ |  |  |
| Emotional Empathy: How well the response demonstrates understanding of the patient's experience |  | ✓ |  |
| Cognitive Empathy: How well each response accounts for the patient's mental state and personal perspective in its recommendations |  | ✓ |  |
| Overall Empathy Score: Overall quality score based on the rater's ranked preference of responses with regard to empathy |  | ✓ |  |
| Overall Readability Score: How easy each response is to understand based on writing style |  |  | ✓ |

## DISCUSSION

The clinical utility of LLMs has been broadly researched in a variety of different medical and otolaryngology-specific contexts^5,13–18^. Our work validates the quality, empathy, and readability of AI-generated responses for a wide array of otologic conditions while also comparing the quality of these responses to that of verified physicians in a blinded setting. Our work continues to add to the body of literature supporting the reliability of these tools but now in a broader otologic context across multiple models^5,13–18^.

Conciseness was notably different between AI-generated and physician responses, which has been observed in prior studies^11,12^. We anticipated that LLMs would generate longer responses, despite being prompted with a word count limit, whereas physicians provided naturally concise replies without guidance. This difference is further influenced by the platform context: Reddit is an informal, conversational environment where physicians tend to offer brief, pragmatic comments rather than comprehensive, clinic-style explanations^19–21^. In contrast, LLMs were explicitly prompted to include supportive language, structured guidance, and clear explanations. Though LLM-generated responses were longer, their length did not compromise their legibility, as ChatGPT still received higher readability scores than physician responses. Nevertheless, response length and readability scores varied between models. Longer responses may be beneficial in this context, as LLM responses were more likely to provide differential diagnoses or medical education than physician responses.

In clinical practice, LLMs may support providers by serving as editable templates that augment, rather than replace, physician involvement in digital patient interactions. These tools may be particularly valuable in providing timely access to medical assistance for individuals who encounter barriers to accessing clinical care. Given our findings and those of the available literature, LLMs harbor the capacity to improve communication quality and responsiveness, and may ultimately strengthen patient engagement and health outcomes^22,23^. This is especially applicable to the context of electronic health record–driven changes to clinical practice.

Increasing reliance on patient portal messaging has expanded physician responsibilities beyond the traditional office visit into non-reimbursable domains and outside of work hours, contributing to clinician burnout and reduced satisfaction^24–26^. LLMs may represent one potential adjunct to reduce this burden while improving patient access to timely information and ultimately decreasing the burden on the healthcare system.

However, as patients increasingly turn to publicly available AI chatbots for guidance without qualified medical input, it is essential to understand the characteristics and communication styles of these tools, as improved empathy and completeness do not substitute for physician judgment^21^. Interestingly, we do note that evaluators were able to identify the response author as AI correctly in 89.4% of cases, which indicates that LLM responses are not yet fully passable for physician responses in this setting. Additionally, LLMs in this study frequently “up-triaged” concerns, recommending high-acuity evaluation even when conservative management might have been reasonable. Medical accuracy still requires verification by trained clinicians, and the expanding technical capabilities of generative AI must be matched with an equally thoughtful understanding of their safe and effective use in healthcare^22^. Despite this, AI may still be used as a supplementary triage mechanism within physician inboxes to help identify potentially concerning messages that may require more urgent review^5^. When appropriately supervised, AI has the potential to enhance the efficiency, clarity, and timeliness of digital communication, allowing clinicians to dedicate more time to direct patient care and in-person clinical encounters^27^.

### Limitations

Several limitations should be noted. First, we acknowledge that digital communication cannot account for the necessary physical examinations or in-office diagnostic procedures that can be fundamental to otologic assessment and clinical decision-making. Even when patients upload images or attempt to describe findings, these inputs do not substitute for direct otoscopic evaluation, limiting the reliability and clinical utility of both online physician and LLM-generated recommendations. In addition, the physician responses analyzed in this study were drawn from an online public forum, where many contributors were not otolaryngologists, did not follow patients longitudinally, and often lacked access to the full clinical context that would typically inform specialist decision-making. The informal and conversational nature of Reddit may also encourage conversational tone, making these responses less representative of professional digital communication provided by healthcare specialists in structured clinical settings.

Although LLM responses were rated as more empathetic, empathy is a multidimensional characteristic that encompasses emotional attunement, relational continuity, and nonverbal communication. These elements may not be fully captured in written text and cannot be reproduced by generative AI to the extent achieved in human interactions. Additionally, LLMs do not routinely provide citations for the information they reference, limiting transparency and making it difficult for patients or clinicians to assess the accuracy or quality of underlying sources. The number of physician raters in this study was small which restricts the generalizability of scoring patterns. Future work should include a larger and more diverse pool of raters, incorporate a broader range of otologic symptoms with varying levels of acuity, and directly compare LLMs with messages written by otologists in secure clinical messaging environments to better evaluate real-world applicability.

## CONCLUSIONS

Our findings suggest that LLMs can generate responses to a wide array of otologic concerns that are highly empathetic, readable, and of comparable quality to those produced by physicians.

Nevertheless, further research is needed to understand how these tools can be responsibly incorporated into clinical workflow, with specific attention to their medical accuracy and safe utilization.

## Data Availability

All data produced in the present study are available upon reasonable request to the authors.

